# Detection of cross-reactive IgA against SARS-CoV-2 spike 1 subunit in saliva

**DOI:** 10.1101/2021.03.29.21253174

**Authors:** Keiichi Tsukinoki, Tatsuo Yamamoto, Keisuke Handa, Mariko Iwamiya, Juri Saruta, Satoshi Ino, Takashi Sakurai

## Abstract

Abundant secretory IgA (sIgA) in mucus, breast milk, and saliva provides immunity that prevents infection of mucosal surfaces. sIgA in pre-pandemic breast milk samples have been reported to cross-react with SARS-CoV-2, but whether it also occurs in saliva and, if so, whether it cross-reacts with SARS-CoV-2, has remained unknown. We aimed to clarify whether sIgA in saliva cross-reacts with SARS-CoV-2 spike 1 subunit in individuals who have not been infected with this virus. The study included 137 (male, n = 101; female, n = 36; mean age, 38.7 [24–65] years) of dentists and doctors in the Kanagawa Dental University Hospital. Saliva and blood samples were analyzed by PCR and immunochromatography for IgG and IgM, respectively. We then identified patients with saliva samples that were confirmed as PCR- and IgM-negative for COVID-19. Proportions of SARS-CoV-2 cross-reactive IgA-positive individuals were determined by enzyme-linked immunosorbent assay using a biotin-labeled spike S1-mFc recombinant protein covering the receptor-binding domain of SARS-CoV-2. The proportion of SARS-CoV-2 cross-reactive IgA-positive individuals was 46.7%, and this correlated negatively with age (r = −0.218, p = 0.01). The proportion of IgA-positive individuals ≥ 50 y was significantly lower than that of patients aged ≤ 49 y (p = 0.008). sIgA was purified from the saliva of all patients, and the salivary sIgA was found to suppress the binding of SARS-CoV-2 spike protein to the ACE-2 receptor. We found SARS-CoV-2 cross-reactive sIgA in the saliva of some participants who had never been infected with the virus, suggesting that sIgA helps prevent SARS-CoV-2 infection.

## Introduction

Secretory IgA (sIgA) prevents infections through mucosal immunity, which is an aspect of the immune system. sIgA, comprising dimeric IgA, a J chain, and a secretory component, is secreted from glandular tissues such as salivary glands and mammary glands onto mucosal surfaces, where it plays a central role in preventing the entry of antigens from the mucosa [1]. Severe acute respiratory syndrome coronavirus 2 (SARS-CoV-2) infects humans via the oral and nasal cavities, and the lungs [2]. The squamous cells of the tongue and periodontal tissues express angiotensin-converting enzyme-2 (ACE-2), a SARS-CoV-2 receptor, transmembrane protease serin 2 (TMPRSS2), and furin, which are proteases that promote infection [3], and saliva can harbor SARS-CoV-2 [4]. Saliva also contains several substances that suppress infection (such as lactoferrin, lysozyme, and sIgA, which is the most abundant) and prevent the virus from entering into the oral cavity [5].

Cross-reactive sIgA (CRsA) against SARS-CoV-2 was identified in breast milk before the COVID-19 pandemic [6, 7]. Furthermore, SARS-CoV-2–reactive CD4+ T cells were also detected in about 40%–60% of unexposed individuals before the pandemic, suggesting that T cells have cross-reactivity to common cold coronaviruses and SARS-CoV-2 [8]. Later findings [9, 10] suggested that prior infection with a coronavirus creates immunological memory that is associated with sIgA cross-reactivity.

Infection with SARS-CoV-2 causes COVID-19, which manifests with a unique spectrum of symptoms ranging from asymptomatic to fatal acute respiratory failure [11]. The severity and prevalence of SARS-CoV-2 infection noticeably differs among age groups and countries, respectively [12]. Immune mechanisms might explain this wide disparity, but they are not yet fully understood. Immunoglobulin G (IgG) can eliminate SARS-CoV-2, and vaccine development against this virus is urgently needed [13]. However, mucosal immunity conferred by sIgA has not been investigated from the viewpoints of recovery from SARS-CoV-2 infection and its prevention. New findings in this area might facilitate deeper understanding of COVID-19 characteristics.

Therefore, we aimed to create an enzyme-linked immunosorbent assay (ELISA) for detecting sIgA that has cross-reactivity with SARS-CoV-2, and revealed whether it has salivary sIgA that cross-reacts with the SARS-CoV-2 spike 1 subunit for non-infected individuals.

## Methods

### Participant selection

We tested saliva and blood samples using PCR and immunochromatography, respectively. Individuals with saliva samples that were confirmed by PCR and IgM testing as negative for COVID-19 were included. The participants included five doctors and 132 dentists at the hospital attached to Kanagawa Dental University. Individuals with IgA nephropathy, selective IgA deficiency, autoimmune diseases, or who had cold-like symptoms within the past 2 weeks were excluded. We analyzed samples from 137 (male, n = 101; female, n = 36; mean age, 38.7 [24–65] years) individuals, under the fully informed consent. This study was conducted under the approval of the Kanagawa Dental University Research Ethics Review Board (Approval number: #690). This study was registered in the Japanese clinical trial UMIN-CTR (Approval number: #R000046461) registry, which meets ICMJE standards.

### Saliva collection for ELISA

We collected samples using Salivettes^®^ (Sarstedt AG & Co. KG, Nümbrecht, Germany) at a fixed room of the hospital between 9 a.m. and 12 p.m. in August 2020, under the infection control. The participants were instructed to refrain from eating, drinking, and brushing their teeth for at least 1 hour before sample collection. The saliva samples were immediately centrifuged at 2,000 × *g* for 15 minutes and then stored at −80°C.

### Design of ELISA for cross-reactive sIgA (CRsA) against spike protein

We modified the ELISA system that could detect IgA cross-reactivity to influenza viruses using human IgA ELISA quantitation set (#E88-102; Bethyl Laboratories, Montgomery, TX, USA) reported by Yamamoto et al. [14]. Saliva samples was diluted 500-fold in carbonate-bicarbonate buffer and incubated for 1 hour at 25°C. Wells were washed five times with wash solution. The antigen used recombinant spike 1-mFc protein (#40591-V05H1; Sino Biological, Beijing, China) comprising the spike 1 subunit with the spike protein receptor-binding domain (RBD) of SARS-CoV-2. The antigen was labeled with biotin using a kit as described by the manufacturer (#BK01; Dojindo Laboratories, Kumamoto, Japan). Biotin-labeled Spike 1 were added to a concentration of 1 µg/mL into wells and incubated for 1 hour at 25°C. Wells were washed five times with wash solution. Next, streptavidin-horseradish peroxidase conjugate (SA202; Millipore, USA; dilution, 1:1000) was used and reacted for 1 hour at 25°C. TMB substrate solution was added for15 mins at 25°C and reaction was quenched with stop solution. Spike 1 protein-bound IgA was determined at 450 nm using a microplate absorbance reader (Bio-Rad Laboratories, Hercules, CA, USA). Background absorbance by the negative control containing PBS was subtracted from the absorbance of all saliva samples.

### IgA purification

We purified sIgA using IgA purification kits (#20395; Thermo Fisher Scientific K.K., Waltham, MA, USA) as described by the manufacturer, then confirmed its molecular weight by standard western blotting using primary anti-human IgA rabbit monoclonal antibodies (ab184863; Abcam Plc, Cambridge, UK) diluted 1:500 and secondary anti-rabbit polyclonal antibodies (#P0448; Dako A/S., Glostrup, Denmark; dilution, 1:1000).

ACE-2 binding to spike protein was inhibited by purified sIgA. We selected and pooled the top 20 antibody-positive, and the bottom 20 antibody-negative samples based on the ELISA results. The final concentrations of antibodies in the pooled positive and negative saliva samples were 93.6 and 63.3 µg/mL, respectively.

### Ability of sIgA antibody to inhibit ACE-2-spike protein binding

The ability of the sIgA to inhibit the binding of ACE-2 to SARS-CoV-2 spike protein was assessed using SARS-CoV-2 spike–ACE-2 binding assay kits (#COV-SACE2-1; RayBiotech, Peachtree Corners, GA, USA), according to the manufacturer. A SARS-CoV-2 spike neutralizing rabbit IgG mAb (#40592-R001; Sino Biological) prepared as a positive control and added neutralizing antibody at concentrations of 0, 0.0125, 0.025, 0.05, 0.1, and 0.2 µg/mL.

### Questionnaire

Participants completed a self-administered questionnaire before saliva collection to determine whether they had previously been inoculated with BCG, hepatitis B, and influenza vaccines within the past year.

### Statistical analysis

Negative values were set to 0 to determine relative ELISA values of CRsA, and association with age was analyzed using Spearman rank correlations. Associations between relative positive (> 0) and negative (0) values of CRsA in ELISA and binary data, age group, gender, BCG vaccination status, hepatitis B vaccination status, and influenza vaccine were examined. Associations with vaccination history were examined using chi-square or Fisher exact tests. We summarized the variables of hepatitis B and influenza vaccines as neither, either, or both to examine associations between the number of vaccinations and IgA positivity or negativity. The variables of BCG, hepatitis B vaccine, and influenza vaccine were summarized as zero, one, two, or three to examine association between the number of vaccinations and IgA positivity or negativity. The significance level was set to 5%. All data were statistically analyzed using SPSS version 26 (IBM Corp., Armonk, NY, USA).

## Results

### Cross-reactive sIgA (CRsA) against spike protein

The relative value of CRsA determined by ELISA was set to 0 as the negative value. The CRsA was positive in 64 (46.7%) and negative in 73 (53.3%) samples.

Figure 2 showed associations between age and cross-reactive IgA. Age significantly and negatively correlated with relative CRsA (r = −0.218, p = 0.01). The positive rate of CRsA was significantly lower in participants aged ≤ 50, compared with those aged ≥ 49 years (p = 0.008). The association between vaccines and CRsA positivity or negativity was not significant.

**Table 1.** Associations between age, sex, and vaccination and CRsA positivity.

| Variables |  | Total | IgA-positive |  | p* |
| --- | --- | --- | --- | --- | --- |
|  |  | n | n | % |  |
| Age (y) | 20-29 | 32 | 19 | 59.4 | 0.081 |
|  | 30-39 | 54 | 27 | 50.0 |  |
|  | 40-49 | 23 | 11 | 47.8 |  |
|  | 50-59 | 20 | 4 | 20.0 |  |
|  | ≥60 | 8 | 3 | 37.5 |  |
|  | 20-49 | 109 | 57 | 52.3 | 0.008 |
|  | ≥50 | 28 | 7 | 25.0 |  |
| Sex | Male | 101 | 48 | 47.5 | 0.452 |
|  | Female | 36 | 16 | 44.4 |  |
| Vaccination |  |  |  |  |  |
| BCG | Yes | 102 | 49 | 48.0 | 0.781 |
|  | No | 8 | 4 | 50.0 |  |
|  | Unknown | 27 | 11 | 40.7 |  |
| Hepatitis B | Yes | 124 | 58 | 46.8 | 0.992 |
|  | No | 11 | 5 | 45.5 |  |
|  | Unknown | 2 | 1 | 50.0 |  |
| Influenza | Yes | 127 | 60 | 47.2 | 0.458 |
|  | No | 10 | 4 | 40.0 |  |
| Hepatitis B<br>and Influenza | Neither | 1 | 1 | 100.0 | 0.114 |
|  | Either | 19 | 7 | 36.8 |  |
|  | Both | 115 | 55 | 47.8 |  |
| BCG,<br>Hepatitis B and<br>Influenza | One | 3 | 1 | 33.3 | 0.809 |
|  | Two | 21 | 11 | 52.4 |  |
|  | Three | 84 | 40 | 47.6 |  |

\*Chi-square or Fisher exact tests.

**Fig. 1.**
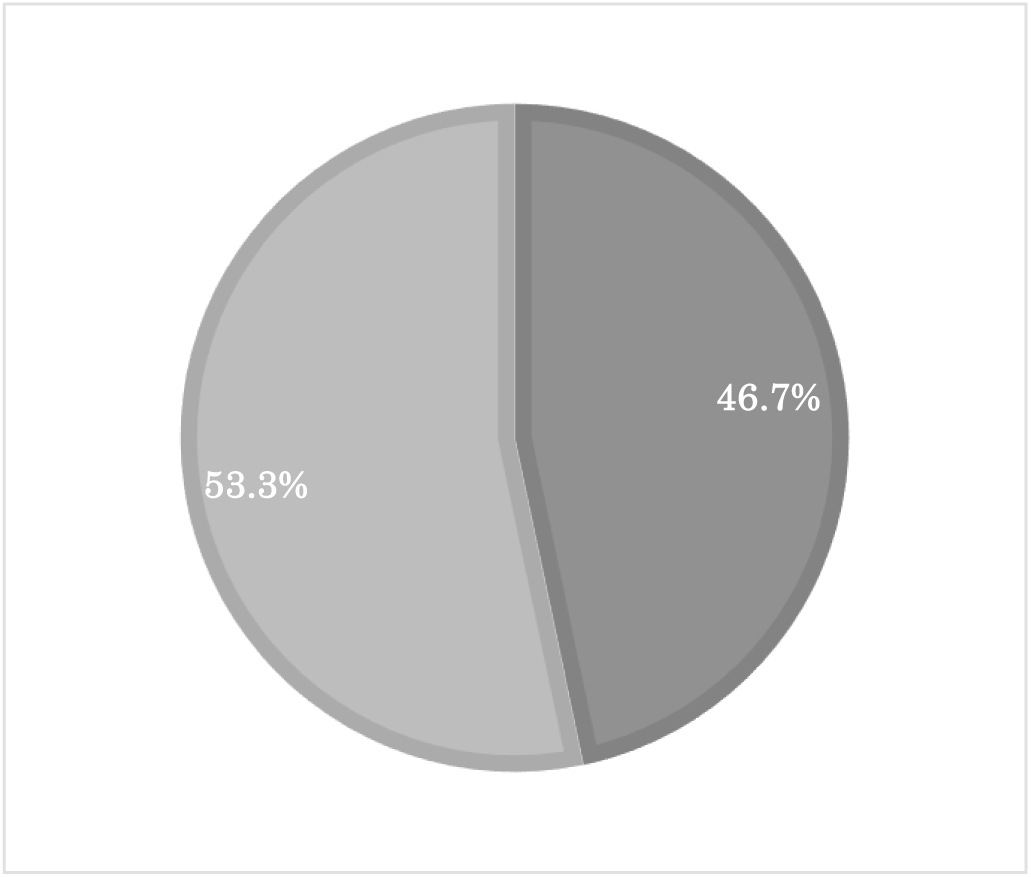
Ratios (%) of detected cross-reactive antibodies to spike protein in saliva. Cross-reactive antibodies in saliva from 137 participants were detected by ELISA. Ratios were calculated as positive or negative (absorbance 0 or less).

**Fig. 2.**
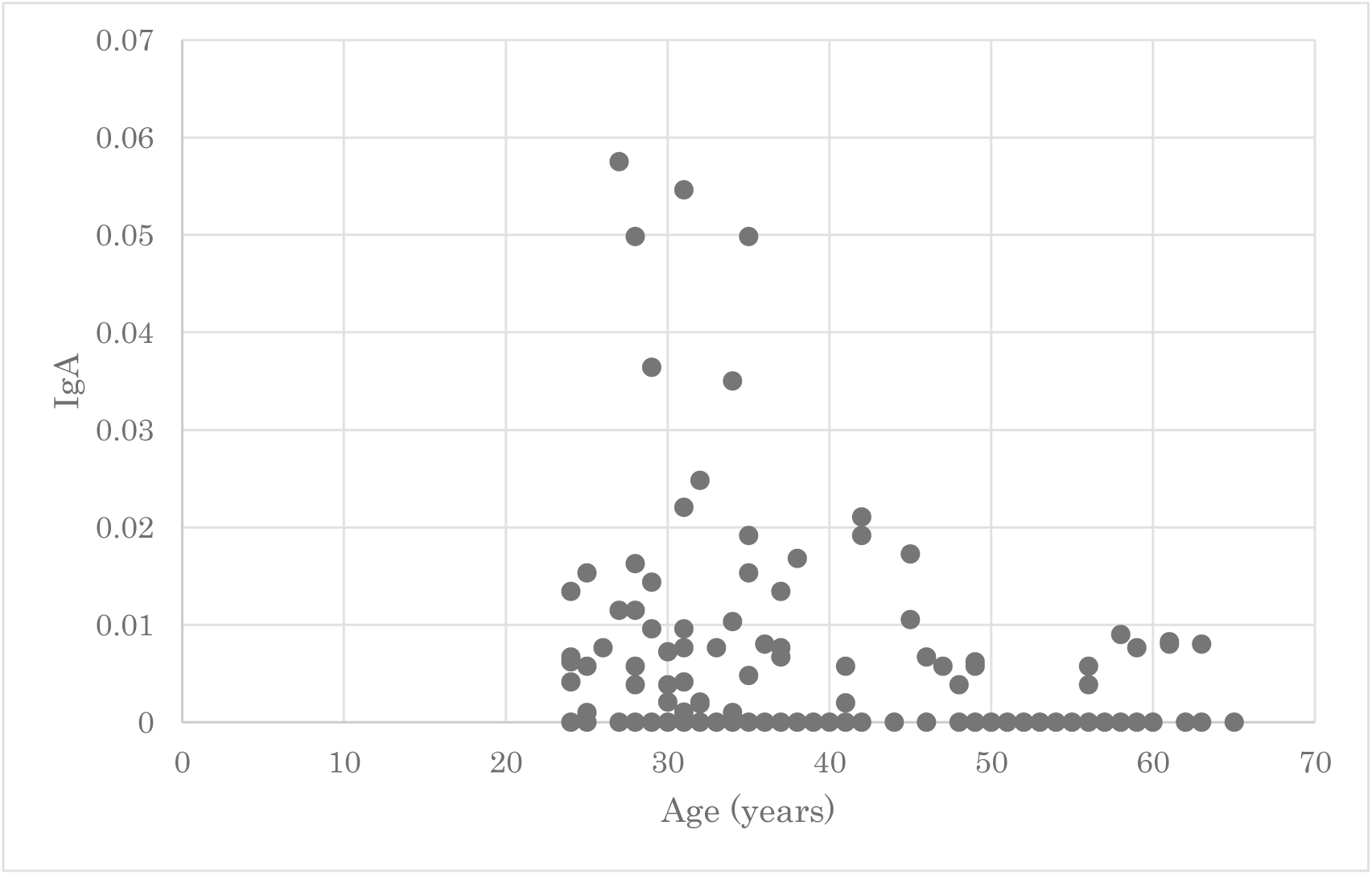
Associations between participant age and positive CRsA. Vertical axis: relative amount of IgA determined as absorbance of CRsA-positive participants. Negative samples are shown as 0. The absorbance was quite high among many participants aged ≤ 49 years.

### CRsA inhibition tests

Western blots revealed purified IgA as a single band of ∼ 60 kDa, confirming that the protein was sIgA (data not shown).

In the positive control, the range of absorbance from the addition of spike neutralizing antibody at different concentrations (0–0.2 µg/mL) was 2.155–0.493, with the concentration-dependent decrease confirming neutralizing activity. The neutralizing antibody against the spike protein showed an absorbance of 2.155 at an antibody concentration of 0 µg/mL. This absorbance indicates the absorbance when ACE-2 and the spike protein are most bound by ELISA. Since the CRsA-negative saliva sample showed an absorbance of 2.402, which was 2.155 or higher, the binding between ACE-2 and the spike protein was not inhibited. The CRsA-positive saliva sample had an absorbance of 1.678, and the binding between ACE-2 and spike protein was partially inhibited.

## Discussion

This study found that 46.7% of saliva samples from 137 participants contained CRsA against SARS-CoV-2 spike 1. The spike 1 region that we examined included the RBD that binds the SARS-CoV-2 receptor ACE-2. This region is important for preventing infections and could thus be a target for vaccine development [14]. A previous ELISA of SARS-CoV-2 CRsA in breast milk found positivity rates of 100% and 80%, respectively, with the total spike protein and RBD as the antigen [6]. However, the salivary values in the present study were lower than both of these. This might be because more sIgA is produced in breast milk than in saliva [15]. The abundance of CRsA in breast milk is a proven post-vaccination response [16], which could explain why the CRsA positivity rate was lower in saliva than breast milk.

Participants with saliva samples that tested negative for COVID-19 determined by PCR and antibody tests did not develop COVID-19 at the time of this article submission. Thus, we believe that they had not been exposed to SARS-CoV-2. As such, we revealed sIgA antibodies against the spike 1 protein of SARS-CoV-2 in individuals with no history of SARS-CoV-2 infection. Only one other study found salivary IgA antibodies that cross-react with the spike protein of SARS-CoV-2 before the COVID-19 pandemic [17]. The nucleocapsid proteins of coronaviruses are highly homologous, but their spike proteins share little commonality [18]. Patients with a history of infection with HCoV-OC43 and other coronaviruses have antibodies against SARS-CoV-1 and antibodies that cross-react to the SARS-CoV nucleocapsid protein, respectively [19]. Furthermore, whereas HCoV-NL63 uses ACE-2 as a receptor, its spike protein shares little homology with SARS-CoV-2 [20]. However, cross-reactivity of the receptor-binding motif 3 of NL63 and COV2-SPIKE_421–434_ of SARS-CoV-2 has recently been reported [20]. As past common coronavirus infection can induce antibodies that are cross-reactive to spike and nucleocapsid proteins, it is reasonable that saliva contains antibodies such as salivary sIgA with cross-reactivity to SARS-CoV-2. However, the epitope behind salivary CRsA cross-reactivity remains to be clarified.

The present study found that CRsA levels decreased with advancing age. This might be explained by the fact that IgA levels decrease with age [21]. In addition, cross-reactive IgG antibodies were identified in 62%, 43.75%, and 5.72% of serum samples from individuals aged 1–16, 17–25, and ≥ 26 years, respectively, before the SARS-CoV-2 pandemic [9]. Because children are infected with common coronaviruses more frequently, this seems to indicate that that more exposure leads to more individuals with cross-reactive antibodies. Moreover, cross-reactive T cells for SARS-CoV-2 are rare in elderly persons [22]. These results can explain why CRsA is prevalent in young persons but uncommon in older persons. The mechanism behind the fact that COVID-19 is less frequently severe and often asymptomatic in children [23] and adolescents [12] might be partially explained.

We showed here that the binding between ACE-2 and spike protein was not inhibited in individuals without CRsA. In contrast, because cross-reactive IgA inhibited the binding of ACE-2 to spike protein, salivary CRsA might prevent SARS-CoV-2 infection *via* a neutralization reaction, because IgA functions as a neutralizing antibody against SARS-CoV-2 [24]. Although saliva contains SARS-CoV-2, it also contains infection inhibitory factors [3]. Saliva lactoferrin is an infection suppressor that binds to SARS-CoV-2 [25]. Because sIgA has an antigen-processing function that is synergistic with lactoferrin, lysozyme, and peroxidase, salivary anti-bacterial or anti-viral factors might enhance the action of CRsA [26].

Early SARS-CoV-2 specific humoral responses are dominated by IgA antibodies, indicating that these play an important role in immunity after SARS-CoV-2 infection [27]. On the other hand, neutralizing IgA antibodies against SARS-CoV-2 persist in saliva for 49 to 73 days after symptoms appear [27]. Spike 1-CRsA in saliva is also associated with the severity of pneumonia in COVID-19 [17]. Saliva can be collected easily and noninvasively, and sIgA is a practical test specimen because it is resistant to degradation and does not have strict transport conditions. Developing a method to easily and non-invasively measure sIgA in saliva might be important for the diagnosis or risk prediction of SARS-CoV-2 infection and responses to vaccines in the future.

One limitation of the present study was the low number of participants. Although vaccines stimulate the production of cross-reactive antibodies [7], we found no significant association between vaccines and cross-reactive antibodies. Future investigations should compare individuals who are not involved in medical care (without experience of vaccination).

## Conclusions

This study identified the SARS-CoV-2 cross-reactive IgA spike protein in saliva from individuals who have not experienced COVID-19. Elderly persons showed lower levels of SARS-CoV-2 spike protein-CRsA than adolescents. Salivary IgA might block the binding of ACE-2 to the spike protein. We revealed the importance of IgA as an inhibitor of SARS-CoV-2 infection in the oral cavity. Our findings are novel, as few studies have examined anti-SARS-CoV-2 antibodies in this compartment, despite the fact of the oral cavity being a viral replication site [28].

## Data Availability

Access to the underlying data is not ready.

## Acknowledgments

We are grateful to Ms. Makiko Yamada at the Central Research Support Center of Kanagawa Dental University Graduate School for providing valuable technical assistance.

## Author contributions

**Conceptualization**: Keiichi Tsukinoki.

**Data analysis**: Tatsuo Yamamoto.

**Sample collection**: Keisuke Handa, Mariko Iwamiya.

**Investigation**: Juri Saruta.

**Project administration**: Satoshi Ino.

**Supervision**: Takashi Sakurai.

**Funding acquisition**: Isamu Kashima.

**Writing-original draft**: Keiichi Tsukinoki.

**Writing-review**: Tatsuo Yamamoto, Keisuke Handa, Juri Saruta.

